# Real-World Effectiveness of a Community-based Multicomponent Maternal Smoking Cessation Program in Preventing Low Birthweight Deliveries: Findings from the CTTP Cohort

**DOI:** 10.1101/2025.08.24.25333399

**Authors:** Syed D. Ahmed, Anne Berit Petersen, Anna P. Nelson, Margarita Martinez, David Shavlik, Bryan Oshiro, Pramil N. Singh

## Abstract

**Introduction:** The effect of smoking cessation during pregnancy on preventing adverse birth outcomes has been shown in studies of US birth certificate data, and in other nations. There is a paucity of data to optimize community-based maternal tobacco cessation programs to improve birth outcomes. Our objective is to evaluate the real-world effectiveness of a multi-component, community-based maternal smoking cessation program in preventing adverse infant outcomes using components of known efficacy.

**Methods:** The Comprehensive Tobacco Treatment Program (CTTP) was a state-funded maternal tobacco smoking cessation program serving pregnant women in San Bernardino County, California, the largest county in the contiguous US. CTTP used a 6-8 week behavioral intervention with components of known efficacy (i.e. incentives, biomarker testing, feedback, and motivational interviewing). We conducted a retrospective cohort study of the 1,402 pregnant women enrolled in CTTP during 2012-2019. We conducted a multivariable logistic regression analysis with adverse infant outcomes (premature birth (PTB), low birthweight (LBW), and NICU admission) as the dependent variables, abstinence achieved during (prolonged abstinence (PA) through weekly urinary cotinine tests) or after the program (self-reported point prevalence abstinence (PPA)) as the main effect exposures, and pertinent confounders.

**Results:** We found that PA during the program significantly decreased the odds of LBW (OR [95% confidence interval (CI) = 0.67 [0.47, 0.96], p = 0.03), and this association remained for self-report of PPA at 2-4 months after the program (OR [95% CI] = 0.70 [0.54, 0.90], p = 0.006), and six months after the program (OR [95% CI] = 0.65 [0.47, 0.90], p = 0.01). Similar, albeit weaker, trends were found for PTB. In these models, older age, early trimester at enrollment, and African American/Black ethnicity also trended toward higher rates of LBW and PTB.

**Conclusions:** Abstinence achieved during a multi-component behavioral smoking cessation intervention program using components of known efficacy significantly reduced low birthweight deliveries in a multi-ethnic population.

## Introduction

Maternal smoking is causally linked to adverse birth outcomes such as preterm birth, low birthweight, small-for-gestational age, neonatal intensive care unit (NICU) admissions, and infant mortality^1^. Mechanisms underlying this effect include fetal hypoxia, toxins in smoke leading to insufficient nutrient availability, teratogenic effects, DNA damage, reduced fetal growth, and an increased risk of congenital abnormalities^2^. Although smoking during pregnancy decreased from 2016 to 2022 in the US^3^, the current rates remain highest among women who are younger, report less than 12 years of education^4^, experience multiple domains of stress before and during gestation^5^, reside in rural areas^6,7^, and/or report indicators of poverty and/or participation in federal assistance programs^5^. By race/ethnicity, smoking during pregnancy is highest in women identifying as American Indian/Alaskan Native, non-Hispanic White, and African American/Black^3^.

Can quitting combustible tobacco smoking during pregnancy prevent or decrease adverse infant outcomes (i.e. preterm birth, low birthweight, NICU admissions, infant mortality)? The association between maternal smoking cessation (self-directed, enrollment in a smoking cessation program) and adverse infant outcomes has been shown in several large cross-sectional studies of US birth certificate data. Using US National Center for Health Statistics data from states recording smoking cessation data on the birth certificate, Soneji et al found that maternal smoking cessation was associated with lower rates of pre-term birth among 25,233,503 expectant mothers^8^. In an analysis of the CDC Pregnancy Risk Assessment and Monitoring System (PRAMS) data from 203,437 birth certificates, Xie et al showed that smoking cessation reduced the prevalence of preterm birth and small-for-gestational age to levels found among non-smokers^5^. These trends linking smoking cessation as a “best buy intervention” to improved birth outcomes are also evident in the global data^9^. There is, however, a paucity of data on optimizing specific components of known efficacy in a maternal smoking cessation for the purpose of improving maternal and child outcomes^10^. A few studies have shown the efficacy of financial incentive-based maternal cessation programs for the prevention of low birth weight.^11^

In this report, we focus on the Comprehensive Tobacco Treatment Program (CTTP) – a state-funded (First 5CA.gov) multi-component smoking cessation program at Loma Linda University Health that served San Bernardino County, California, during 2012-2019. San Bernardino County is, by land area, the largest county in the US with a multi-ethnic population of over two million; infant mortality rates in the county have long followed health disparity trends by factors such as race/ethnicity and poverty^12^. The programmatic approach of CTTP was a behavioral intervention (about 8 weeks) delivered in a group format (classroom setting) by health educators, and used intervention components of known efficacy (i.e., incentives, biomarker testing, feedback, and motivational interviewing). During a program evaluation of CTTP in 2020, the data from all 1,402 participants were analyzed as a retrospective cohort study that we have described in detail^13^. The overall aim of the present study is to examine whether achieving abstinence during or after the multi-component CTTP behavioral intervention decreased the rate of selected adverse infant outcomes at delivery (preterm birth (PTB), low birthweight (LBW), NICU admissions).

## Methods

The methods to assemble and analyze the CTTP cohort data have been extensively described by Petersen et al^13^. Here, we briefly summarize the cohort, the outcomes, main effect exposures, confounder variables, and statistical methods.

### CTTP Cohort

All program participants from 2012-2019 were enrolled in the CTTP cohort (n = 1,402). Participants were recruited from a county-wide referral network maintained by health educators at CTTP. Referral sites included outpatient clinics, hospitals, and rehabilitation facilities. Tobacco use was screened according to standardized prenatal protocols^14^. Self-enrollment was also available and publicized via flyers located at WIC offices, community events, and health fairs.

### CTTP Multicomponent Behavioral Smoking Cessation Program

The program was a six-to-eight-week smoking cessation intervention for pregnant women residing in San Bernardino County. The program incorporated weekly in-person sessions that included cotinine-verified abstinence testing, motivational interviewing, personalized quit plans, and education on the risks of smoking tobacco. Incentives such as diapers and xylitol gum were provided for each cotinine-negative week to encourage adherence. Health educators delivered individualized counseling using the ACOG 5 A’s framework, along with screening for depression, alcohol, and substance use, with referrals as needed^15^. Follow-up for all participants occurred through telephone appointments at 2-4 months and 6 months after the program administration. This telephone appointment was also used to assess current abstinence and offer relapse support if necessary.

### CTTP Outcomes

Dependent variables include PTB, LBW, NICU admission, and a combined adverse outcome variable. PTB was classified as a birth occurring before 37 weeks of gestation^16^. As per World Health Organization guidelines (WHO), LBW was defined as a neonate weighing less than 2,500 grams at birth^17^. NICU admission was assessed by the health educators during follow-up appointments after program administration. The combined adverse outcome was computed as a composite variable indicating the occurrence of any one of the adverse outcomes (PTB, LBW, NICU admission).

### CTTP Main Effect Variables and Pertinent Confounders

A main effect exposure variable for prolonged abstinence (PA) during the behavioral intervention program was defined as completing the program with six to eight consecutive weeks of cotinine-verified abstinence assessed as urinary cotinine. After the program administration, all subjects (completers, non-completers) were offered a telephone appointment support during which point prevalence abstinence (PPA) from tobacco was assessed by self-report. PPA was assessed at 2-4 months and 6 months after program administration and is also used as a main effect exposure variable.

Pertinent confounders include self-reported (by interview) age at enrollment, trimester at enrollment, number of cigarettes smoked at enrollment, and race/ethnicity. Household smoking was measured from a self-reported (by participant interview) list of household members (i.e. spouse, partner, family member, roommate) who smoke cigarettes.

### Statistical Methods

Descriptive statistics of the cohort have been published previously^13^ and, in this report, we provide a cohort profile by level of adverse birth outcome. Prevalences and means are given with asymptotic 95% confidence intervals. Differences by levels of adverse birth outcome are assessed by contingency table methods (i.e. chi-squared) or, for continuous variables, by independent sample t-tests.

The relation between maternal smoking cessation and adverse infant outcomes was assessed in logistic regression models. In each of the models an abstinence measure (PA achieved at program completion, PPA at 2-4 months after the program, PPA at 6 months after the program) was the main effect, and the dependent variables were adverse infant outcomes (PTB, LBW, NICU admission). Pertinent confounders were tested by a change of estimate approach^18^. Model fit was tested using the Hosmer-Lemeshow test for continuous variables and log likelihood ratio test for indicator variables^18^. Missing values were imputed using well established methods for multiple imputation from Rubin^19^. All statistical analyses, including multiple imputation^20^, were conducted using SAS 9.4 (Cary, NC). The Loma Linda University Health Institutional Review Board approved the secondary analysis study protocol (IRB # 5190418).

## Results

Descriptive statistics of the CTTP cohort (n = 1,402) have been previously reported.^13^ Briefly, the mean age of the pregnant women of the cohort is 26.8 (standard deviation(SD)=5.8), the mean gestational weeks at delivery was 38.8 (SD=3.2), and the most common race/ethnicity was Hispanic/Latina (42.9%). Also, we have previously reported that 40.1% of the cohort achieved prolonged abstinence(PA) as defined by testing negative (urinary cotinine) during each week of their enrollment in a completed program offering of the CTTP.^21^ In Table 1, we have reported pertinent characteristics of the cohort by subgroup of experiencing adverse birth outcomes (low birthweight (LBW), pre-term birth (PTB), admission of the neonate to the neonatal intensive care unit (NICU). These data show no substantial differences by age at enrollment and cigarettes smoked at enrollment. Adverse birth outcomes were more prevalent in women who enrolled in CTTP during the first and second trimesters, and also more prevalent in African American/Black women.

**Table 1.** Selected characteristics of the Comprehensive Tobacco Treatment Program Cohort (n = 1,402) enrolled during 2012-2019 are given by birth outcome category.

|  |  |  |  |
| --- | --- | --- | --- |
| Variable | Adverse Birth Outcomes |  | p value |
|  | Yes | No |  |
| **Age at Enrollment (years)** | 27.8 | 27.4 | 0.30 |
| **Number of Cigarettes Smoked at Enrollment** | 1.6 | 1.4 | 0.33 |
| **Trimester at Enrollment (%)** |  |  | 0.04 |
| First | 7.4 | 5.6 |  |
| Second | 36.6 | 27.6 |  |
| Third | 56 | 66.8 |  |
| **Race/Ethnicity (%)** |  |  |  |
| African American/Black | 22.3 | 16 | 0.02 |
| White | 16.6 | 28.5 |  |
| Hispanic/Latino | 53.7 | 46.6 |  |
| Native American/Alaskan Native | 1.1 | 0.4 |  |
| Asian/Pacific Islander | 1.7 | 2.5 |  |
| More than One Ethnicity | 4.6 | 5.9 |  |
| **Household Smoking (%)** |  |  |  |
| Yes | 53.0 | 47.0 | 0.29 |
| No | 49.0 | 51.0 |  |

We conducted multivariable logistic regression analyses with 1) specific adverse birth outcomes (LBW, PTB, NICU admissions) as the outcome variable, 2) abstinence during (PA) or after the program (point prevalence abstinence (PPA) at 2-4 months after the program, PPA at 6 months after the program) as the main effect exposure variables, and 3) confounders for age at enrollment, cigarettes smoked at enrollment, trimester at enrollment, and race/ethnicity. We note that associations between PA and birth outcomes are longitudinal since CTTP completion occurred during gestation. The odds ratios linking PPA at 2-4 months and 6 months to birth outcomes are more cross-sectional in nature since these measures often occurred after delivery.

In Table 2, we provide the findings from these models where LBW or PTB was the outcome variable. We found a significant decrease in the odds of LBW for PA (OR [95% confidence interval (CI)] = 0.67 [0.47, 0.96]), PPA at 2-4 months (OR [95% CI] = 0.70 [0.54, 0.90]), and PPA at 6 months (OR [95% CI] = 0.65 [0.47, 0.90]). In these models, third-trimester enrollments in CTTP were about 30% less likely to result in a LBW delivery, and African American/Black mothers were two-fold more likely to experience a LBW delivery. Similar albeit weaker trends were found linking PA and PPA to PTB in Table 2. For NICU admission (not shown), no strong or significant association was found with PA or PPA.

**Table 2.** Multivariable Logistic Regression Models for Low Birth Weight and Pre-term Birth Outcomes, an Abstinence Main Effect, and Four confounders from the Comprehensive Tobacco Treatment Program Cohort (n = 1402).

|  | **OR [95% CI] for Low Birth Weight** |  |  |  | **OR [95% CI] for Pre-Term Birth** |  |  |
| --- | --- | --- | --- | --- | --- | --- | --- |
| **Variables in Model** | **Model 1** | **Model 2** | **Model 3** |  | **Model 1** | **Model 2** | **Model 3** |
| **Main Effect (Abstinence):** |  |  |  |  |  |  |  |
| Prolonged Abstinence during the Program† | 0.67 [0.47, 0.96]* |  |  |  | 0.80 [0.63, 1.01] |  |  |
| Point Prevalence Abstinence 2-4 months after Program‡ |  | 0.70 [0.54, 0.90]** |  |  |  | 0.80 [0.63, 1.03] |  |
| Point Prevalence Abstinence 6 months after Program‡ |  |  | 0.65 [0.47, 0.90]** |  |  |  | 0.80 [0.62, 1.02] |
| **Confounders:** |  |  |  |  |  |  |  |
| Age at Enrollment | 1.02 [0.97, 1.08] | 1.03 [0.99, 1.07] | 1.03 [0.99, 1.07] |  | 1.06 [1.02, 1.11]** | 1.07 [1.02, 1.11]*** | 1.07 [1.03, 1.11]** |
| Number of Cigarettes Smoked at Enrollment | 1.03 [0.93, 1.13] | 1.03 [0.96, 1.11] | 1.04 [0.97, 1.12] |  | 0.99 [0.94, 1.05] | 0.99 [0.93, 1.05] | 1.00 [0.94, 1.06] |
| Trimester at Enrollment |  |  |  |  |  |  |  |
| First | 1.08 [0.70, 1.67] | 1.07 [0.74, 1.53] | 0.70 [0.36, 1.33] |  | 1.31 [1.00, 1.72]* | 1.30 [0.99, 1.70] | 1.31 [0.94, 1.82] |
| Second | 1.0 [referent] | 1.0 [referent] | 1.0 [referent] |  | 1.0 [referent] | 1.0 [referent] | 1.0 [referent] |
| Third | 0.75 [0.58, 0.98]* | 0.78 [0.61, 1.01] | 0.79 [0.62, 1.02]* |  | 0.61 [0.50, 0.75]**** | 0.61 [0.50, 0.76]*** | 0.62 [0.51, 0.76]**** |
| Race/Ethnicity |  |  |  |  |  |  |  |
| African American/Black | 1.27 [0.93, 1.74] | 1.26 [0.91, 1.73] | 1.27 [0.92, 1.73] |  | 1.27 [0.98, 1.64] | 1.30 [0.99, 1.71] | 1.31 [0.99, 1.72] |
| White | 1.0 [referent] | 1.0 [referent] | 1.0 [referent] |  | 1.0 [referent] | 1.0 [referent] | 1.0 [referent] |
| Hispanic/Latino | 1.13 [0.81, 1.59] | 1.14 [0.82, 1.57] | 1.11 [0.80, 1.55] |  | 1.22 [0.95, 1.56] | 1.26 [1.00, 1.59]* | 1.25 [0.98, 1.59] |
| Native American | 1.36 [0.50, 3.70] | 1.36 [0.48, 3.89] | 1.42 [0.49, 4.05] |  | - | - | - |
| Asian/Pacific Islander | - | - | - |  | 0.65 [0.24, 1.73] | 0.72 [0.27, 1.95] | 0.72 [0.26, 1.96] |
| More than One Ethnicity | 1.46 [0.85, 2.49] | 1.45 [0.87, 2.42] | 1.47 [0.88, 2.45] |  | 0.98 [0.63, 1.51] | 0.97 [0.61, 1.55] | 0.98 [0.62, 1.55] |
| Household member(s) Smoking |  |  |  |  |  |  |  |
| Yes | 1.21 [0.95, 1.55] | 1.20 [0.96, 1.52] | 1.20 [0.95, 1.52] |  | 1.13 [0.96, 1.34] | 1.13 [0.94, 1.36] | 1.14 [0.95, 1.37] |
| No | 1.0 [referent] | 1.0 [referent] | 1.0 [referent] |  | 1.0 [referent] | 1.0 [referent] | 1.0 [referent] |

\* $p \leq 0.05$ , \*\* $p \leq 0.01$ , \*\*\* $p \leq 0.001$ , \*\*\*\* $p \leq 0.0001$
† Testing negative for urinary cotinine during each week of enrollment in the completed program
± Self-report of not smoking during the past seven days.
- insufficient subjects to compute a point estimate

We also ran models (not shown) where we combined all birth outcomes into an Adverse Birth Outcome (LBW, PTB, or NICU admission) dependent variable. We found no significant associations for PA or PPA.

## Discussion

Our evaluation of the CTTP cohort indicated that prolonged abstinence (PA) – achieved by completing weekly negative cotinine tests throughout the program administration – significantly decreased the odds (OR [95% confidence interval (CI)] = 0.67 [0.47, 0.96], p = 0.03) of low birthweight (LBW), and this association remained for point prevalence abstinence (PPA) at 2-4 months after the program (OR [95% CI] = 0.70 [0.54, 0.90], p = 0.006), and six months after the program (OR [95% CI] = 0.65 [0.47, 0.90], p = 0.01). Slightly weaker (ORs of about 0.8) and non-significant associations were found between the abstinence measures (PA, PPA) and preterm birth (PTB), and the association with NICU admissions was close to the null.

Taken together, our findings show that a multi-component intervention for pregnant women who smoke cigarettes can significantly decrease low birthweight deliveries. The CTTP intervention uses components of known efficacy, such as financial incentives^11,22^, biomarker testing^23^, biofeedback^22^, and motivational interviewing^22^. Also, as shown in a Cochrane review of effective maternal smoking cessation interventions, CTTP used a high frequency of counseling sessions tailored to maternal smoking cessation: 6-8 weekly contacts (1 hour of a group class) and telephone follow-up that continued post-partum.^22^ Concordant with the multi-component CTTP approach, the Cochrane review concludes that the best results are achieved by combining financial incentives with more frequent counseling sessions^22^.

### Preventing Low Birthweight and Pre-term Birth through Smoking Cessation: The San Bernardino County Experience

The CTTP cohort provides insight into the efficacy of a multi-component maternal smoking cessation program in a multi-ethnic county (52% Hispanic/Latino, 24% White, 9% Asian, 9% African/American Black, 4%More than one race, <1% American Indian/Alaskan Native, <1% Pacific Islander). This is important since smoking during pregnancy disproportionately affects American Indian/Alaskan Native, non-Hispanic White, and African/American communities.

Our group has previously reported that an analysis of the birth certificate data from San Bernardino County indicated that for every 35 pregnant women who quit cigarette smoking, one pre-term birth was prevented^24^. From these data, we estimated that half of the pregnant smokers in the county who did not quit smoking during pregnancy on their own enrolled in CTTP^24^ – indicating excellent outreach. Moreover, a 40.1% abstinence rate was achieved by CTTP^21^. Taken together, our findings from these analyses indicate that the CTTP approach of combining excellent outreach with intervention components of known efficacy can significantly reduce important adverse birth outcomes such as low birth weight.

Our findings from CTTP are concordant with at least one study that used a similar multi-component approach to maternal smoking cessation and related abstinence to adverse birth outcomes. The “Baby and Me Tobacco Free” (BMTF) was first designed and implemented in New York in 2011 and since then has been implemented in 21 states^25^. BMTF incorporates components of financial incentives, biofeedback through carbon monoxide testing, and motivational interviewing (four sessions). In Colorado, BMTF involved a data linkage with the PRAMS study and thus was able to relate abstinence achieved in the program to adverse birth outcomes. Among 2,231 participants in BMTF, Polinski et al found that BMTF enrollees had a significantly lower risk of PTB, LBW, and NICU admissions relative to the 16,739 pregnant smokers in the control sample from PRAMS who did not enroll in BMTF.^11^

### Limitations

A number of limitations of this analysis of CTTP data need to be discussed. The CTTP cohort may not have had statistical power to detect effects with all birth outcomes (PTB, LBW, NICU admissions). This may explain why we only found a significant association with LBW as compared to the BMTF analyses of over eighteen thousand women^11^. In CTTP we have previously reported that despite having excellent outreach, we did have a high rate of dropout^13^. We have posited that one reason for the dropout rate is the travel time to a program run in a fixed classroom setting. Our current work involves adding a home visit/tele visit approach to the CTTP model. Also, our birth outcomes are self-reported, and this may introduce bias. Lastly, it is important to note that since CTTP data are from 2012-2019, we did not capture sufficient exposure to e-cigarettes or legalized cannabis, which are more prevalent among pregnant women today.^26^ E-cigarette and cannabis use patterns (exclusive, dual user) need consideration in the design of future interventions.

### Conclusion

Our findings from a multi-ethnic sample of 1,402 pregnant women who smoked cigarettes during pregnancy, it indicate that abstinence achieved during a multi-component maternal smoking cessation program using components of known efficacy significantly reduced low birthweight. We were able to control for important confounders in the analysis (age, nicotine dependence at enrollment, trimester at enrollment, race/ethnicity, and household members who smoke) and demonstrate the impact of single-site maternal smoking cessation program serving the largest county in the US. Our findings need confirmation in larger prospective samples that also consider current e-cigarette and cannabis exposure among pregnant women in these communities.

## Data Availability

All data produced in the present study are available upon reasonable request to the authors.

